# External validation of a treatment decision algorithm for tuberculosis in children living with HIV - a diagnostic cohort study

**DOI:** 10.1101/2024.11.08.24316648

**Authors:** Celso Khosa, Minh Huyen Ton Nu Nguyet, Juliet Mwanga-Amumpaire, Chishala Chabala, Raoul Moh, Clementine Roucher, Denis Nansera, Bwendo Nduna, Eugenia Macassa, Madeleine Amorrissany Folquet, Dalila Rego, Gae Mundundu, Naome Natukunda, Perfect Shankalala, Saniata Cumbe, Eric Komena, Andrew P. Steenhoff, Anneke C. Hesseling, James A Seddon, Eric Wobudeya, Maryline Bonnet, Olivier Marcy, TB-Speed HIV study group

## Abstract

**Introduction:** Tuberculosis (TB) is the leading cause of death in children living with HIV (CLHIV) and is challenging to confirm the diagnosis. The PAANTHER treatment decision algorithm (TDA) was developed to improve the diagnosis of TB in CLHIV. We aimed to externally validate the PAANTHER TDA in CLHIV with presumptive TB.

**Methods:** We conducted a prospective diagnostic cohort study in seven tertiary hospitals across Côte d’Ivoire, Mozambique, Uganda, and Zambia, implementing the PAANTHER TDA in CLHIV aged between 1 month and 14 years with presumptive TB. TDA assessments included Xpert MTB/RIF Ultra (Ultra) on respiratory and stool samples, history of contact, symptoms (fever >2 weeks, unremitting cough, haemoptysis and/or weight loss in previous 4 weeks, tachycardia), chest radiography and abdominal ultrasound. A positive score (≥100) prompted TB treatment initiation. Children were followed-up for 6 months, and retrospectively classified as having confirmed, unconfirmed or unlikely TB. The primary outcome was the proportion of missed TB cases (false negative) among children with negative scores; secondary outcomes included TDA diagnostic accuracy, feasibility, and time to treatment initiation. The TDA was considered validated if the negative predictive value (NPV, 1 - rate of false negative) was superior to a 75% pre-established confidence interval lower limit.

**Findings:** From 2 October 2019 to 31 December 2021, we enrolled 277 CLHIV, including 175 (63·2%) who were on antiretroviral therapy at inclusion. 272 (98·2%) children had a complete TDA evaluation; 215 (75.8%) scored >100, including 24 (8·7%) with positive Ultra. 182 (86·7%) children who scored ≥100, and 12 children who scored negative, initiated TB treatment at a median of 1 (IQR: 0-3) and 27 [8·2; 64] days after inclusion, respectively. 62/215 children (28·8%) who scored ≥100 were classified as having unlikely TB and 4/12 (33·3%) who scored negative were initiated on treatment and were classified as having unconfirmed TB. The proportion of children with TB (confirmed and unconfirmed) was 155/273 (56·8%; 95% CI: 50·9; 62·5). The NPV was 55/67 (93·3%; 95% CI: 84·1; 97·4), reaching protocol-defined validation. The TDA sensitivity was 97·4% (95% CI: 93·6; 90·0) with specificity of 47·5 (95% CI: 38·7; 56·4).

**Interpretation:** The PAANTHER TDA was validated in CLHIV. Its high sensitivity, excellent feasibility, and short turnaround time to treatment initiation, should allow rapid treatment decision-making and could reduce morbidity and mortality in CLHIV.

**Funding:** UNITAID

## INTRODUCTION

Tuberculosis (TB) remains an important public health concern, particularly among children living with HIV (CLHIV). According to the World Health Organization (WHO), in 2022, of the 1.3 million TB-related deaths estimated worldwide, 183,000 were in children aged 0-14 years old, and 31,000 were in CLHIV.^1^ Most of the death are assumed to be in children never initiated on antituberculosis treatment Tuberculosis is the leading cause of death, and the leading opportunistic infection, in this vulnerable population. TB poses diagnostic challenges in CLHIV due in part to the low sensitivity of diagnostic tests, driven by the paucibacillary presentation of the disease, and in part by the poorly specific and atypical symptoms caused by the underlying immunodeficiency.^2^

Undiagnosed and untreated TB increases the risk of death in children, especially in CLHIV, who are highly vulnerable to developing severe TB disease. Symptom-based approaches that perform well in HIV-negative children may perform poorly in CLHIVHIV.^3^ Despite innovations in disease management and treatment, accurate TB diagnosis in CLHIV continues to be challenging, necessitating an alternative approach and novel tools. Rapid diagnosis and treatment are crucial due to more severe clinical presentation, drug-drug interactions, and higher TB mortality rates.^2^ Hospital-based studies report case-fatality ratios of 14–41% in CLHIV receiving TB treatment.^4–6^ A retrospective study in Cape Town found that CLHIVHIV, treated for TB, had higher mortality and twice the odds of an unfavourable outcome compared to HIV-negative children.^7^

Due to impaired immunological containment of *M. tuberculosis*, tuberculosis disease progresses more rapidly in CLHIV, compared to HIV-negative children, and leads to more severe disease. Untreated, tuberculosis mortality is higher in CLHIV, compared to HIV-negative children and even if diagnosed and treated, mortality is higher for CLHIV.^3^ This is in part due to more severe disease, in part due to underlying immunosuppression, and in part due to delayed diagnosis, resulting from poorly specific symptoms, signs and radiology. Hospital-based studies report case-fatality ratios of 14–41% in CLHIV receiving TB treatment.^4–6^ A retrospective study in Cape Town found that children living with HIV, treated for TB, had higher mortality and twice the odds of an unfavourable outcome compared to HIV-negative children.^7^ To reduce mortality from tuberculosis in CLHIV, it is necessary to identify more CLHIV who have tuberculosis and to diagnose them earlier, before the disease has become severe. Symptom-based approaches that perform well in HIV-negative children may perform poorly in children living with HIV,^3^ and despite access to molecular diagnosis and antigen based point-of-care tests^8^, accurate tuberculosis diagnosis in CLHIV continues to be challenging, necessitating an alternative approach.

The PAANTHER treatment decision algorithm (TDA) was developed using data collected from CLHIV evaluated for tuberculosis in Cambodia, Vietnam, Cameroon and Burkina Faso^9^. The clinical, microbiological and radiological characteristics from these children were evaluated for their ability to discriminate children with tuberculosis from those with other diseases, and the strength of that discriminatory power scaled into a scoring system that could be used to make treatment decisions. When evaluated internally, the PAANTHER scoring system performed with a sensitivity of 88.6% and specificity of 61.2%.^9^ External validation, however, is a critical component for evaluating the effectiveness and generalizability of TDAs. There are currently no published studies describing the external evaluation of TDA performance in diverse clinical, geographic, and epidemiological settings, particularly in low- and middle-income settings (LMICs).

Given this gap, we sought to externally validate the PAANTHER TDA in CLHIV with presumptive TB by assessing its diagnostic performance, feasibility, and impact on treatment decisions in diverse clinical, geographic and epidemiologic settings.

## METHODS

### Study design and participants

We carried out a prospective diagnostic cohort study in seven tertiary hospitals in Côte d’Ivoire, Mozambique, Uganda and Zambia. These four countries have high TB incidence and HIV prevalence. We assessed the overall diagnostic performance and feasibility of the PAANTHER TDA, and the safety of withholding TB treatment in those with a negative PAANTHER TB score. The key validation criterium was based on the occurrence of algorithm failures of children with missed TB (i.e. false negatives).

From 2 October 2019 to 31 December 2021, we enrolled children aged 1 month to 14 years with documented HIV-infection who had presumptive TB based on at least one of the following criteria: i) persistent cough for more than 2 weeks, ii) persistent fever for more than 2 weeks, iii) failure to thrive in the last three months, iv) failure of broad-spectrum antibiotics for the treatment of pneumonia, v) chest X-ray (CXR) features suggestive of tuberculosis. Also included were children with a history of tuberculosis exposure combined with any of the symptoms listed above with a shorter duration (< 2 weeks). We excluded children who were receiving TB treatment at the time of screening or in the preceding 3 months, except for children on TB preventive treatment. Written informed consent was obtained from parents or guardians, and assent from children aged ≥7.

The study (NCT04121026) was approved by the sponsor’s (Inserm) institutional ethics review committee, the WHO ethics review committee, as well as the national ethics committees and institutional review boards in Côte d’Ivoire, Mozambique, Uganda and Zambia (see appendix).

### Procedures

Children with presumptive TB were evaluated by study nurses following identification at outpatient clinics and inpatient wards, having screened positive for tuberculosis based on WHO symptom-based criteria by attending routine care clinicians and nurses. Eligible children were invited to participate to the study after obtaining consent from the parents/guardians and assent from the children, in compliance with local regulations.

As per the PAANTHER TDA, at inclusion, children underwent: i) clinical evaluation for a history of close exposure to an adult with bacteriologically confirmed tuberculosis and assessment for suggestive tuberculosis symptoms (prolonged fever for more than 2 weeks, unremitting cough, haemoptysis and/or weight loss in the past 4 weeks and tachycardia, ii) microbiological testing using the molecular Xpert MTB/RIF Ultra (Xpert; Cepheid, Sunnyvale, CA, USA) assay on one sample each of a nasopharyngeal aspirate, a stool sample and either two expectorated sputa samples or gastric aspirates, iii) a chest radiograph, and iv) abdominal ultrasonography to assess for the presence of intra-abdominal lymphadenopathy.

In addition, for the purpose of the study, additional clinical signs and symptoms and ultrasonography features were recorded. Mycobacterial culture was performed on two expectorated sputa or gastric aspirates, and children had blood sampling for additional laboratory tests (see supplementary methods). All data were collected using an electronic case report form developed in REDCap, accessible on remote devices. This case report form also had a score calculation module. The algorithm and its score were additionally available in paper format. attending routine care clinicians were encouraged to initiate tuberculosis treatment without delay based on the PAANTHER TDA score (see Appendix Figure 1). The diagnostic process was expected to last one or two days. Tuberculosis treatment using at standard 6 months WHO-recommend treatment was initiated in children with a score ≥100. Flexibility to start tuberculosis treatment in children with a score <100 was allowed in cases of severe/life threatening conditions, at the clinician’s discretion.

All children had a diagnostic visit at 1-2 days 2 following inclusion and were seen on day 15 and at months 1, 2, 3 and 6. For children not initiated on TB treatment at baseline, a visit was planned at day 7 to identify those with possible missed TB. At each visit, children had a medical history taken, underwent a clinical evaluation for any new symptoms or signs of tuberculosis and to evaluate for adverse events, adherence to tuberculosis medications, nutritional recovery, and symptom resolution. A chest radiograph was performed, and blood was taken for laboratory tests at months 2 and 6. Specimen collection with Ultra testing and chest radiography could be repeated if a child without a tuberculosis diagnosis, showed no clinical improvement or presented with new symptoms or signs of tuberculosis at any time. The tuberculosis treatment outcomes were assessed at 6 months.

### Case review committees

At the end of the study, children were retrospectively classified as having confirmed, unconfirmed or unlikely tuberculosis. The classification process differed between those with negative and those with positive PAANTHER scores with an endpoint review committee (ERC) established in each country and a centralized international ERC made of independent expert not involved in the management of the study. Each country ERC reviewed: i) children with clinically diagnosed tuberculosis with a negative score; ii) differential diagnoses in children not clinically improving at month 2; iii) all deaths and ongoing disease at the time of death for causality. Children with negative scores that had subsequently been initiated on tuberculosis treatment were classified using the updated NIH Clinical Case Definition for Classification of Childhood Intrathoracic Tuberculosis^10^ by the country ERC. Children identified as false negatives by national ERCs were reviewed and the final classification was attributed by the centralized international ERC. Children with positive PAANTHER scores without microbiological confirmation were classified as either unconfirmed or unlikely tuberculosis based on expert opinion by the centralized international ERC. This was because the use of the updated Clinical Case Definitions for Classification of Intrathoracic Tuberculosis in Children could not be used as the decision to start treatment because it was not clinician-based but algorithm-guided.

During the COVID-19 pandemic lockdowns in the study countries (April to October 2020), follow-up was conducted by phone when physical visits were not possible. The safety of staff and children/families were ensured at the study visits, and at times of sample collection and sample processing in the laboratory.

An Independent Data Monitoring Committee monitored the safety of the study through regular safety reports.

### Outcomes

The primary outcome was the proportion of children with missed tuberculosis (false negatives) in children not initiated on treatment as per the PAANTHER TDA. Secondary outcomes included: a) time to final tuberculosis treatment decision, b) proportion of children with presumptive tuberculosis having completed the PAANTHER TDA (i.e. either positive or with tuberculosis ruled out based on the score), c) proportion of children considered as unlikely tuberculosis by the ERC in those initiated on treatment as per the PAANTHER TDA (false positives), and d) diagnostic accuracy (negative and positive predictive values and corresponding sensitivity and specificity) of the PAANTHER TDA overall and in the subgroups of age, immunodepression, nutritional, and antiretroviral therapy status.

### Statistical analysis

Based on a hypothesized tuberculosis prevalence of 50% and an estimated TDA sensitivity and specificity of 89% and 61%,^9^ respectively, we estimated the proportion of missed tuberculosis cases among children not started on treatment based on the PAANTHER algorithm (assessed by tuberculosis incident cases in the 6-month period after applying the algorithm) to be 15%. This corresponds to a negative predictive value (NPV) of 85%. For sample size calculations, we used an unacceptable NPV of 75% (minimal acceptable 95% lower confidence interval limit), a one-sided test of level 5% and a probability β=5% (power=95%), which resulted in a sample of 176 children not initiated on treatment as per the PAANTHER TDA and a total sample of 550 children, taking into account an expected 10% of missing data.^11^ Following discussions with the Scientific Advisory Board on feasibility and budgetary issues, study recruitment was concluded on December 2021, when 277 participants had been enrolled.

We evaluated the proportion of missed tuberculosis in CLHIV with presumptive tuberculosis not initiated on treatment as per the PAANTHER TDA (i.e. false negatives), and estimated the NPV of the PAANTHER TDA and its 95% confidence interval (CI). We compared the lower limit of the 95%CI to the minimal acceptable lower confidence interval limit to assess whether the algorithm was validated. We assessed the proportion of over-diagnosed tuberculosis among children with a negative PAANTHER score (i.e. false positive), and estimated the positive predictive value (PPV) of the PAANTHER TDA, tuberculosis prevalence in the study, the sensitivity and specificity of TDA, and their 95% CIs. We assessed and compared the diagnostic accuracy of the TDA between subgroups. *Post-hoc*, we assessed the concordance between items as scored by clinicians and clinical information reported in the case report forms using Cohen’s Kappa statistic and we carried out a sensitivity analysis comparing the diagnostic performance of the PAANTHER TDA as scored by the clinician with that of the TDA score recalculated *a posteriori* using clinical information entered in the case report form.

### Role of the funding source

The study funder (UNITAID) had no role in the study design, data collection, data analysis, data interpretation, or writing of the report.

## RESULTS

Of 713 CLHIV screened for tuberculosis symptoms, 423 were eligible and 277 were enrolled in the study (Figure 1). The median (interquartile range [IQR)]) age was 5·0 (2·0; 10·0) years, 142 (51·6%) were female, 175 (70·0%) were on antiretroviral therapy at recruitment, median CD4 count was 736 (325·8; 1,302) cells/µl, median CD4 percentage was 21·0 (IQR: 10·1; 30·0), and 84 (41·8%) children had severe immunosuppression (Table 1). Of children recruited, 172 (62·8%) children had a persistent cough for more than 2 weeks, 90 (32·8%) had a persistent fever for more than 2 weeks, 177 (64·6%) were failing to thrive, 61 (22·3%) had failed broad-spectrum antibiotics for the treatment of pneumonia, 189 (72·4%) had features on chest radiography suggestive of tuberculosis, and 18 (6·6%) had history of exposure to an adult with bacteriologically confirmed tuberculosis, together with symptoms of a shorter duration.

**Figure 1.**
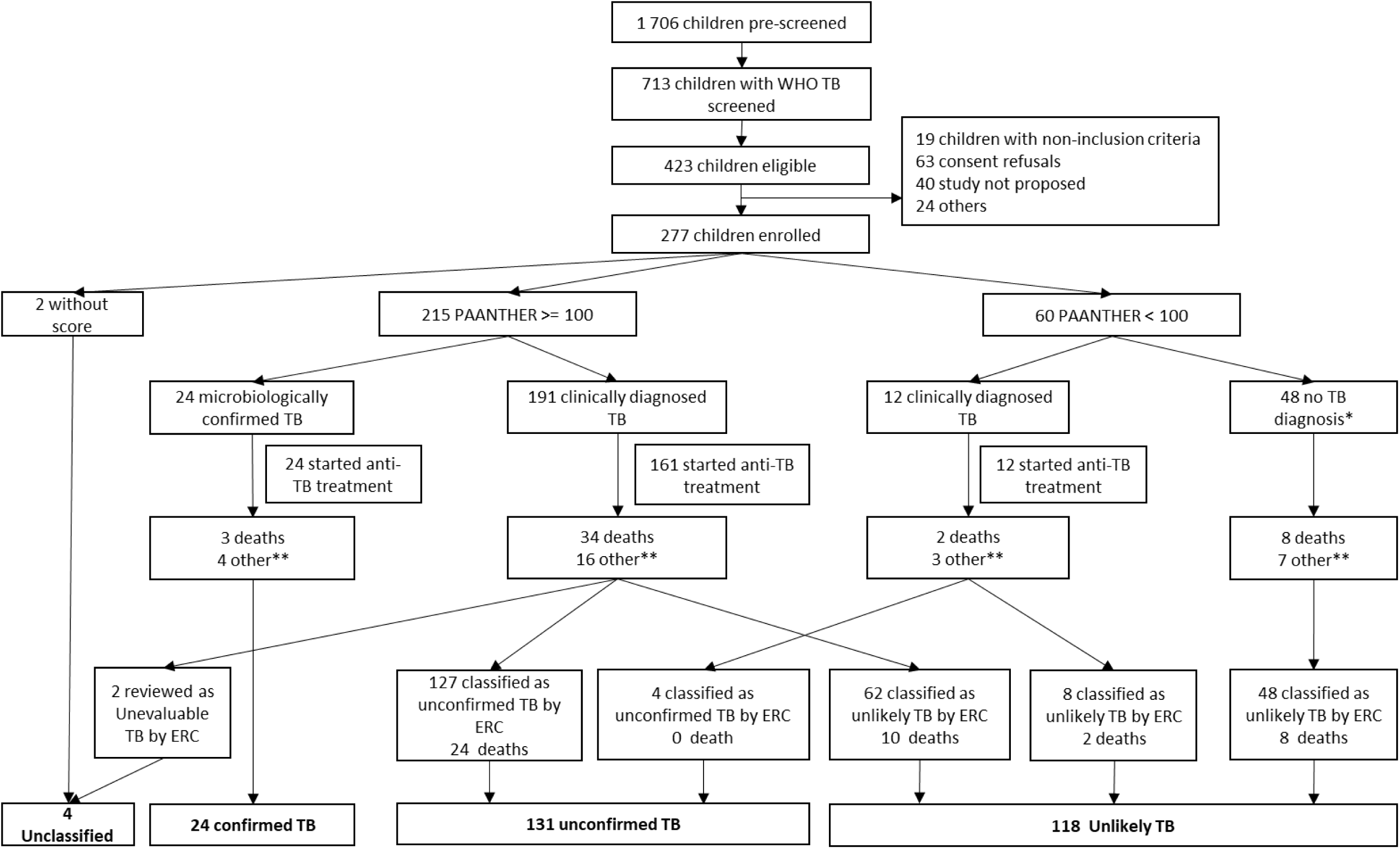
Study flow chart. TB: tuberculosis; ERC: endpoint review committee. *One child with score <100, no TB diagnosis or treatment, with trace positive Xpert MTB/RIF Ultra result nasopharyngeal aspirate, alive; ** Other = Loss-to-follow-up, Transfer, Withdrawal.

**Table 1.** Children with a calculated score characteristics. All data are n (%) or median IQR).

| Characteristic | N' if different from N | All children N = 275 n (%) or median [IQR] | N' if different from N | Positive score N = 215 n (%) or median [IQR] | N' if different from N | Negative score N = 60 n (%) or median [IQR] | P-value |
| --- | --- | --- | --- | --- | --- | --- | --- |
| Age (years) |  | 5 (2·0; 10) |  | 5 (2·0; 10) |  | 5 [2·1; 10] | 0·98 |
| Sex female |  | 142 (51·6) |  | 110 (51·2) |  | 32 (53·3) | 0·77 |
| On ART at inclusion | 250 | 175 (70·0) | 198 | 132 (66·7) | 52 | 43 (82·7) | 0·025 |
| CD4 count (cells/mm3) | 208 | 736 (325·8; 1 302) | 165 | 730 (330·0; 1 283) | 43 | 870 (312·5; 1 424) | 0·51 |
| CD4 percentage | 206 | 21 [10·1; 30] | 164 | 21 [10·0; 29] | 42 | 20 [13·4; 30] | 0·94 |
| Severe immunosuppression<sup>a</sup> | 201 | 84 (41·8) | 160 | 66 (41·2) | 41 | 18 (43·9) | 0·43 |
| Weight for Age Z-score | 201 | -3 (-4·1; -1) | 153 | -3 (-4·0; -1) | 45 | -3 (-4·3; -1) | 0·37 |
| Severe Acute Malnutrition<sup>b</sup> |  | 118 (42·9%) |  | 93 (43·3%) |  | 25 (41·7%) | 0·83 |
| BCG vaccination | 249 | 194 (70·5) | 192 | 150 (69·8) | 57 | 44 (73·3) | 0·41 |
| Previous TB treatment |  | 19 (6·9) |  | 14 (6·5) |  | 5 (8·3) | 0·74 |
| Exposure to person with smear or Xpert + tuberculosis <sup>c</sup> | 274 | 18 (6·6%) |  | 17 (7·9%) | 59 | 1 (1·7) | 0·13 |
| Pulmonary infection in the past month |  | 132 (48·0) |  | 117 (54·4) |  | 15 (25·0) | 0·47 |
| Failure of broad-spectrum antibiotics for pneumonia | 274 | 61 (22·3) | 214 | 51 (23·8) |  | 10 (16·7) | 0·24 |
| Cough (any, ongoing) | 274 | 237 (86·5) |  | 196 (91·2) | 59 | 41 (69·5) | <0·0001 |
| Cough > 2 weeks | 274 | 172 (62·8) |  | 151 (70·2) | 59 | 21 (35·6) | <0·0001 |
| Unremitting cough<sup>c</sup> | 274 | 136 (49·6) |  | 119 (55·3) | 59 | 17 (28·8) | 0·0003 |
| Fever (any, ongoing) | 274 | 161 (58·8) |  | 138 (64·2) | 59 | 23 (39·0) | 0·0005 |
| Fever > 2 weeks<sup>c</sup> | 274 | 90 (32·8) |  | 84 (39·1) | 59 | 6 (10·2) | <0·0001 |
| Haemoptysis in previous 4 weeks<sup>c</sup> | 274 | 2 (0·7) |  | 2 (0·9) | 59 | 0 (0·0) | >0·99 |
| Weight loss in previous 4 weeks<sup>c,d</sup> | 274 | 177 (64·6) |  | 142 (66·0) | 59 | 35 (59·3) | 0·34 |
| Loss of appetite | 274 | 143 (52·2) |  | 117 (54·4) | 59 | 26 (44·1) | 0·16 |
| Fatigue or loss of playfulness | 274 | 140 (51·1) |  | 115 (53·5) | 59 | 25 (42·4) | 0·13 |
| Tachycardia<sup>c</sup> |  | 62 (22·5) |  | 53 (24·7) |  | 9 (15·0) | 0·11 |
| Cervical lymphadenopathy | 274 | 63 (23·0) |  | 55 (25·6) | 59 | 8 (13·6) | 0·052 |
| Hepatomegaly | 274 | 47 (17·2) |  | 42 (19·5) | 59 | 5 (8·5) | 0·046 |
| Chest radiography features consistent with tuberculosis | 261 | 189 (72·4) | 205 | 176 (85·9) | 56 | 13 (23·2) | <0·0001 |
| Miliary<sup>c</sup> |  | 32 (12·3) |  | 31 (15·1) |  | 1 (1·8) | 0·0070 |
| Alveolar opacity<sup>c</sup> |  | 136 (52.1) |  | 126 (61.5) |  | 10 (17.9) | <0.0001 |
| Lymph nodes<sup>c</sup> |  | 126 (48.3) |  | 124 (60.5) |  | 2 (3.6) | <0.0001 |
| Cavitation |  | 25 (9.6) |  | 23 (11.2) |  | 2 (3.6) | 0.085 |
| Pleural effusion |  | 13 (5.0) |  | 12 (5.9) |  | 1 (1.8) | 0.31 |
| Airway compression |  | 4 (1.5) |  | 4 (2.0) |  | 0 (0.0) | 0.58 |
| Abdominal Lymph nodes<sup>c</sup> | 258 | 36 (14.0) | 204 | 35 (17.2) | 54 | 1 (1.9) | 0.0039 |
| Xpert MTB/RIF Ultra positive<sup>c</sup> | 271 | 25 (9.2) | 212 | 24 (11.3) | 59 | 1 (1.7)<sup>e</sup> | 0.024 |
| MTB culture positive on any sample | 228 | 8 (3.5) | 179 | 8 (4.5) | 49 | 0 (0.0) | 0.21 |
| PAANTHER total score |  | 208 (118.5; 285) |  | 242 (179.0; 303) |  | 63 (39.0; 90) | <0.0001 |
| TB treatment initiated |  | 197 (71.6) |  | 185 (86.0) |  | 12 (20.0) | <0.0001 |
| Reference tuberculosis classification | 273 |  | 213 |  |  |  | <0.0001 |
| Confirmed |  | 25 (9.2) |  | 24 (11.3) |  | 1 (1.7) |  |
| Unconfirmed |  | 135 (49.5) |  | 127 (59.6) |  | 8 (13.3) |  |
| Unlikely |  | 113 (41.4) |  | 62 (29.1) |  | 51 (85.0) |  |
| End of study status |  |  |  |  |  |  | 0.24 |
| Alive |  | 198 (72.0) |  | 158 (73.5) |  | 40 (66.7) |  |
| Dead |  | 47 (17.1) |  | 37 (17.2) |  | 10 (16.7) |  |
| Transferred out |  | 1 (0.4) |  | 0 (0.0) |  | 1 (1.7) |  |
| Withdrawn |  | 12 (4.4) |  | 9 (4.2) |  | 3 (5.0) |  |
| Lost to follow-up |  | 17 (6.2) |  | 11 (5.1) |  | 6 (10.0) |  |

Overall, 275 (99·3%) children had a score calculated by the clinician and 272 (98·9%) had a complete score (. Around 90% had Xpert testing on microbiological samples, as well as abdominal ultrasound performed (Table 2), and 97·1% had chest radiography performed. All tests were completed within a median time of 1 day. 215 (78·2%) children were scored positive, including 24 (8·7%) with microbiological confirmation, and 60 (21·8%) were scored negative by the clinicians (Figure 1). The clinician scoring and the supporting data in the study case report form were concordant in 248 (90·2%) cases overall on the total score (Cohen’s kappa = 0·73 [0·63; 0·83]), with higher discrepancies on tuberculosis exposure history, unremitting cough and tachycardia (supplementary appendix table 3S3).

**Table 2.** TB assessments and score items, feasibility (score and scoring items uptake), time to score items and overall score, time to final TB treatment decision, time to start of TB treatment.

|  | **Feasibility**<br>**(uptake)**<br>n (%) | **Time to score**<br>**items (days)**<br>median [IQR] |
| --- | --- | --- |
| Complete scoring<sup>a</sup> | 272 (98.9) | NA |
| Xpert Ultra on nasopharyngeal aspirates | 257 (92.8) | 1 [0; 1] |
| Xpert Ultra on stools and/or expectorated sputum | 248 (89.5) | 1 [1; 2] |
| Chest radiography | 269 (97.1) | 0 [0; 1] |
| Abdominal ultrasound | 259 (93.6) | 1 [0; 2] |
| TB treatment initiation | 197 (71.1) | 1 [0; 4] |

<sup>a</sup>scoring was considered as complete if total PAANTHER score was at least 100, or if all score items were assessed if total PAANTHER score was below 100.

Tuberculosis treatment was initiated in 197 (71·1%) children at a median time of 1 (0; 4) day after inclusion, including 185/215 (86·0%) children with a positive score, that were initiated within a median of 1 (IQR: 0; 3) day, and 12/60 children with a negative score that initiated treatment at a median time of 27 (IQR: 8·2; 64) days after inclusion. Overall, 47 (17·1%) children died during the study, including 37/215 (17·2%) children with a positive score and 10/60 (16·7%) children with a negative score. In those 30 children with a positive score who were not initiated on tuberculosis treatment, 5 (16·7%) children died, including two on the day of enrolment, one on the 31^st^, one on the 55^th^ and one on the 87^th^ days after enrolment.

After ERC review, 62/215 (28·8%) children with a positive score were classified as unlikely tuberculosis, with 2 as unevaluable. Four out of 12 children who had a negative score but who had been started on tuberculosis treatment were classified as unconfirmed tuberculosis (Figure 1). The resulting tuberculosis proportion in the classifiable study population (N=273) was 56·8% (95%CI: 50·9; 62·5) with 131 unconfirmed and 24 confirmed tuberculosis.

The score calculated by the clinician had a NPV of 93·3 (95%CI: 84·1; 97·4), therefore reaching the per protocol validation criteria. The score had a sensitivity of 97·4 (95%CI: 93·6; 99.0) and a specificity of 47·5 (95%CI: 38·7; 56·4) to detect tuberculosis ((Table 3). The subgroup analyses showed significant differences in the specificity and the PPV according to the presence/absence of severe immunosuppression, and countries (Table 3).

**Table 3.**
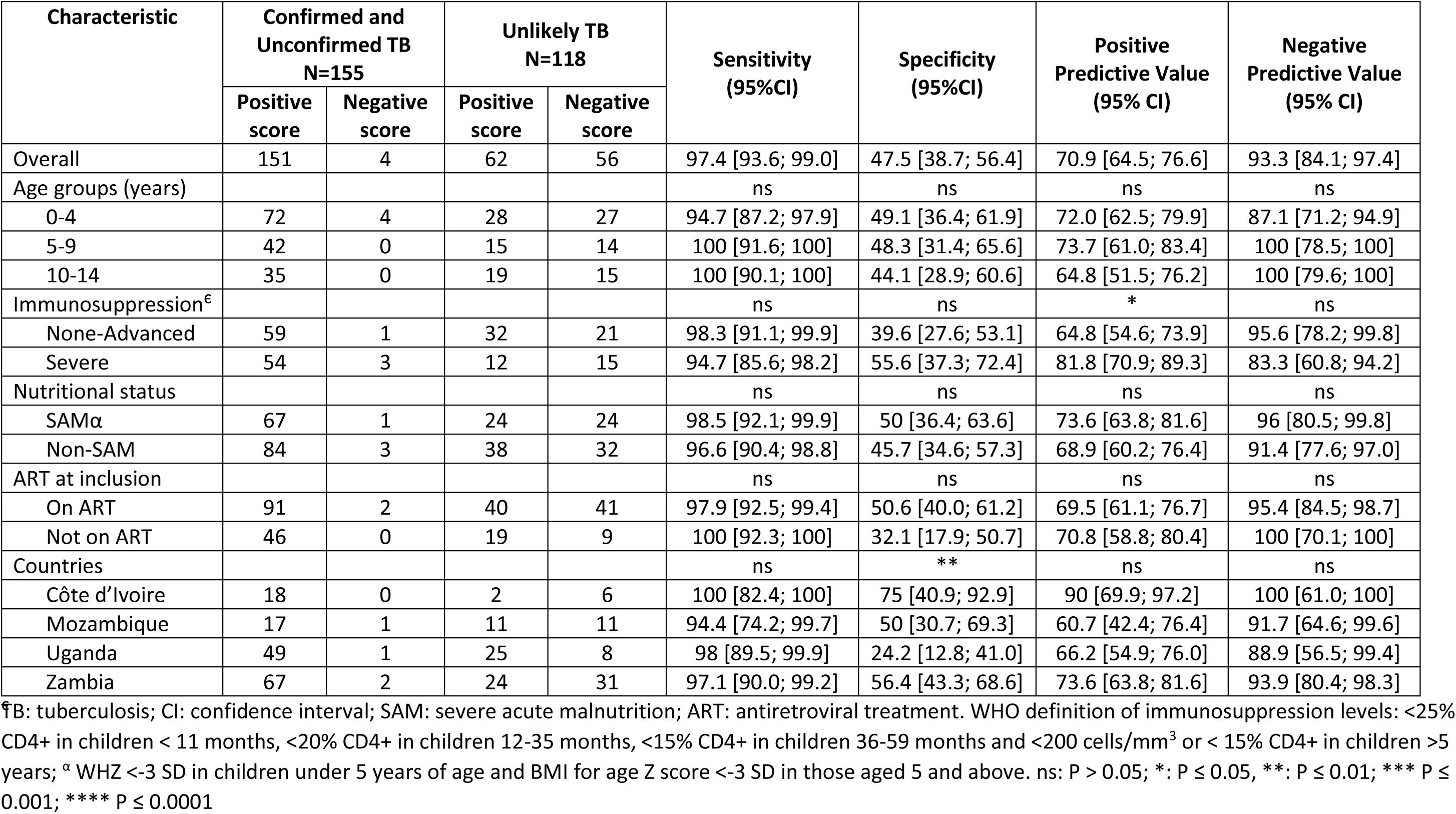
PAANTHER score overall diagnostic performance in the cohort and in subgroups.

The diagnostic performance of the score, recalculated based on clinical features reported in the case report forms, differed significantly from that of the clinician calculated scoring on sensitivity (McNemar P-Value < 0.05, see supplementary appendix table S4) but not on specificity (McNemar P-Value = 0.999).

## DISCUSSION

We validated the PAANTHER TDA for tuberculosis diagnosis in CLHIV, showing that its NPV reached the protocol-defined validation criteria. The TDA showed high feasibility and enabled short turnaround time to results and treatment initiation. This is a key step to support implementation of this diagnostic tool in CLHIV in high tuberculosis and HIV burden settings.

Overall, there were very few tuberculosis diagnoses missed by the PAANTHER TDA, showing that sensitivity is maintained when implementing it in other settings. This fits with the TDA development strategy that favoured sensitivity, aiming to reduce mortality in this vulnerable group.^12^ Clinicians using the TDA made a treatment decision within a median time of one day. Short time to treatment initiation is essential to reduce mortality.^13^ In the development study, the median time to tuberculosis diagnosis was 7 days and the delay is therefore substantially reduced, as was hypothesised during TDA development.^9^ There were no deaths in children with negative scores classified as unconfirmed tuberculosis in this validation study. The specificity was lower than estimated previously, which could have several explanations. First, the unremitting cough criteria was assessed using a graphic method,^14^ which was reportedly challenging to use by clinicians; that may have contributed to over-diagnosing cough lasting more than 2 weeks as unremitting cough, therefore contributing to a decreased specificity and increased sensitivity. Second, the reference classification in children with a positive score could not rely on positive response to treatment as the treatment decision was forced by the TDA; this may have led to biased estimates of the tuberculosis prevalence and the diagnostic accuracy. At last, the specificity was much lower in one study country, which could be explained by more clinicians from this setting over-diagnosing chronic cough as unremitting. Our results, however, suggest that, for CLHIV, the PAANTHER algorithm may outperform, in terms of diagnostic accuracy, the WHO-suggested TDA A (the algorithm that includes chest radiography) which proposes a single scoring approach for the general paediatric population, not specific for HIV-positive or -negative children with an estimated sensitivity of 86% and a specificity of 37%, for the symptom and chest radiography scoring part.^15,16^

To our knowledge, this study is the first prospective external validation of a data-driven TDA. Since the publication of the PAANTHER algorithm, several data-driven TDAs have been developed, in HIV-negative children,^17^ in the general paediatric population,^15^ in children hospitalized with severe acute malnutrition,^18^ and other TDAs have been developed for use when molecular testing is unavailable.^19,20^ None of these algorithms have been externally validated to date. We prospectively implemented TDAs for real life treatment decision-making, and therefore faced the methodological issues inherent to childhood tuberculosis diagnosis research, mostly critically in relation to the lack of an independent reference standard. Indeed, due to the treatment decision enforced by the TDA, the evolution under or off treatment, which is a strong contributor to classify children as either unconfirmed or unlikely tuberculosis,^21^ could not be used as a classifying criteria for the reference standard.^10^ Despite challenges, and although more thorough implementation research on use of TDAs is needed, this type of external validation study, even at tertiary levels of care, is closer to real life implementation than estimates of the diagnostic accuracy of TDAs, using data already collected.^15^ Lessons learned included that a positive score did not automatically lead to tuberculosis treatment initiation. This was notably true in one of the settings where treatment initiation was made by another program team that did not necessarily follow the clinician decision based on TDA scoring. Our study also highlighted challenges in accurate reporting of clinical findings in the scoring system which calls for more user-friendly digital tools for use of TDAs.

Despite its good performance, implementing the PAANTHER TDA at lower levels of healthcare may face challenges. One of the challenges expected will be the availability of both radiography and ultrasonography in decentralized settings. Of note, due to the low specificity of clinical features, both investigations contributed to improvements in the specificity of the TDA development models and derived scores by 7-20%.^15,22^ A better specificity is important in vulnerable children, such as CLHIV, as in addition to tuberculosis, it is important to diagnose and treat promptly other conditions.

Implementing the PAANTHER TDA alongside WHO-recommended TDAs for other children may be challenging for health professionals. Clinical decision support systems, enabling the use of multiple TDAs, could contribute to alleviating the cognitive load on the physician, and to move toward more individualised approaches in global health. In addition to supporting diagnostics, these systems could support decision-making around the type of treatment needed, an important consideration with the recent introduction of a shorter treatment regimen for less severe tuberculosis in children.^23,24^

This study has limitations. First, due to the COVID-19 pandemic, logistics, and budgetary issues, we recruited a smaller sample than planned initially. Although we were able to reach the TDA validation criteria per protocol, this smaller sample may have biased our results and contributed to less precise estimates. Second, we validated the tool in tertiary referral hospitals, while it is intended to be implemented mostly at secondary levels of healthcare. This may have positively impacted the feasibility, turnaround time to results, and TDA diagnostic accuracy as compared to routine conditions at lower levels of healthcare. Third, we did not perform qualitative assessments of acceptability and feasibility from the clinician’s perspective, two aspects that are considered essential in the evidence requested by WHO in order to inform the GRADE process for guideline development.^25^ Fourth, as mentioned above, the classification process for the reference standard may have contributed to biased estimates of the prevalence, hence of the diagnostic accuracy of the PAANTHER TDA. Lastly, we did not collect data on the screening step, hence are unable to determine how this algorithm integrates with the WHO-recommended 4 question screening approach that targets acute symptoms.^26^

Despite improved access to antiretroviral therapy, increases in tuberculosis screening through the WHO 4 question strategy, and access to urine lipoarabinomannan testing point of care, CLHIV remain at high risk of developing tuberculosis, high risk of being undiagnosed for the disease, and at high risk of tuberculosis-related mortality. The PAANTHER TDA specifically developed and now validated for CLHIV with presumptive tuberculosis aims to guide clinicians in a standardised process for improved diagnosis and subsequent access to treatment. This study provides the first evidence for external validation of TDAs as requested by WHO, and demonstrated that the PAANTHER TDA has potential for a substantial public health impact through wide scale use in CLHIV by HIV and tuberculosis programs. Given its current performance with a high sensitivity above the WHO target for diagnostic tests, and still sub-optimal specificity, the PAANTHER TDA could still be improved secondarily by the addition of biomarkers, such as urine lipoarabinomannan or a host-response immunological biomarker, or simple score recalibration and adjustments. Artificial intelligence read radiography or point of care ultrasonography systems could additionally make these approaches more widely available. Validating these new versions of the TDA may warrant novel approaches to external validation to shorten the time for evidence generation, allowing more rapid transfer of scientific evidence into policy and practices, enabling public health impact on tuberculosis-related mortality in children.

## Data Availability

All data produced in the present work are contained in the manuscript

## Acknowledgements

Preliminary findings from this study were presented at the Union Conference 2022. Inserm (Pôle Recherche Clinique), Paris, France was the sponsor of the trial. We thank the Ministries of Health and National Tuberculosis programs of participating countries for their support. We thank the members of the independent data monitoring committee, Muriel Rabilloud (chair, Hospices Civils de Lyon, Lyon, France), Mohammod Jobayer Chisti (Dhaka Hospital, Dhaka, Bangladesh), and Helena Rabbie (Tygerberg Hospital, Stellenbosch University, Cape Town, South Africa), and the members of the TB-Speed Scientific Advisory Board who gave technical advice on the design of the study and approved the protocol: Steve Graham (chair, University of Melbourne, Melbourne, Australia), Anneke Hesseling (Stellenbosch University, Cape Town, South Africa), Luis Cuevas (Liverpool School of Tropical Medicine, UK), Christophe Delacourt (Hôpital Necker-Enfants Malades, France), Sabine Verkuijl (WHO, Switzerland), Philippa Musoke (Makerere University, Uganda), Mark Nicol (University of Western Australia, Perth, Australia), Elizabeth Maleche-Obimbo (University of Nairobi, Kenya), as well as Chishala Chabala (University of Zambia) and Mao Tan Eang (CENAT, Cambodia) who represented other TB-Speed investigators at Scientific Advisory Board meetings. We thank all the children and their families who participated in the trial, and the healthcare workers of the participating hospitals and laboratories.

## Conflict of interest declarations

We declare no competing interests.

## Author contributions

OM, MB and EW conceived and designed the study and led the study at the international level. CR coordinated study implementation at the international level while NN, PS, SC, and EK coordinated practical study implementation in Uganda, Zambia, Mozambique, and Côte d’Ivoire respectively, under scientific leadership from JMA, CC, CK, and RM, respectively. AH provided scientific guidance and expertise for the study, through the Scientific Advisory Board. JMA, CC, RM, DN, BN, EM, MAF, DR, and GM implemented the study and enrolled participants. JAS and APS classified children as members of the international expert review committee. MN performed the statistical analysis. CK, OM, MH, MB and EW contributed to the interpretation of the results. CK wrote the first draft, and all authors reviewed and approved the final version of the report and the manuscript. CK, OM, MB, and EW were responsible for the decision to submit the manuscript.

## SUPPLEMENTARY APPENDIX

### Ethics committees

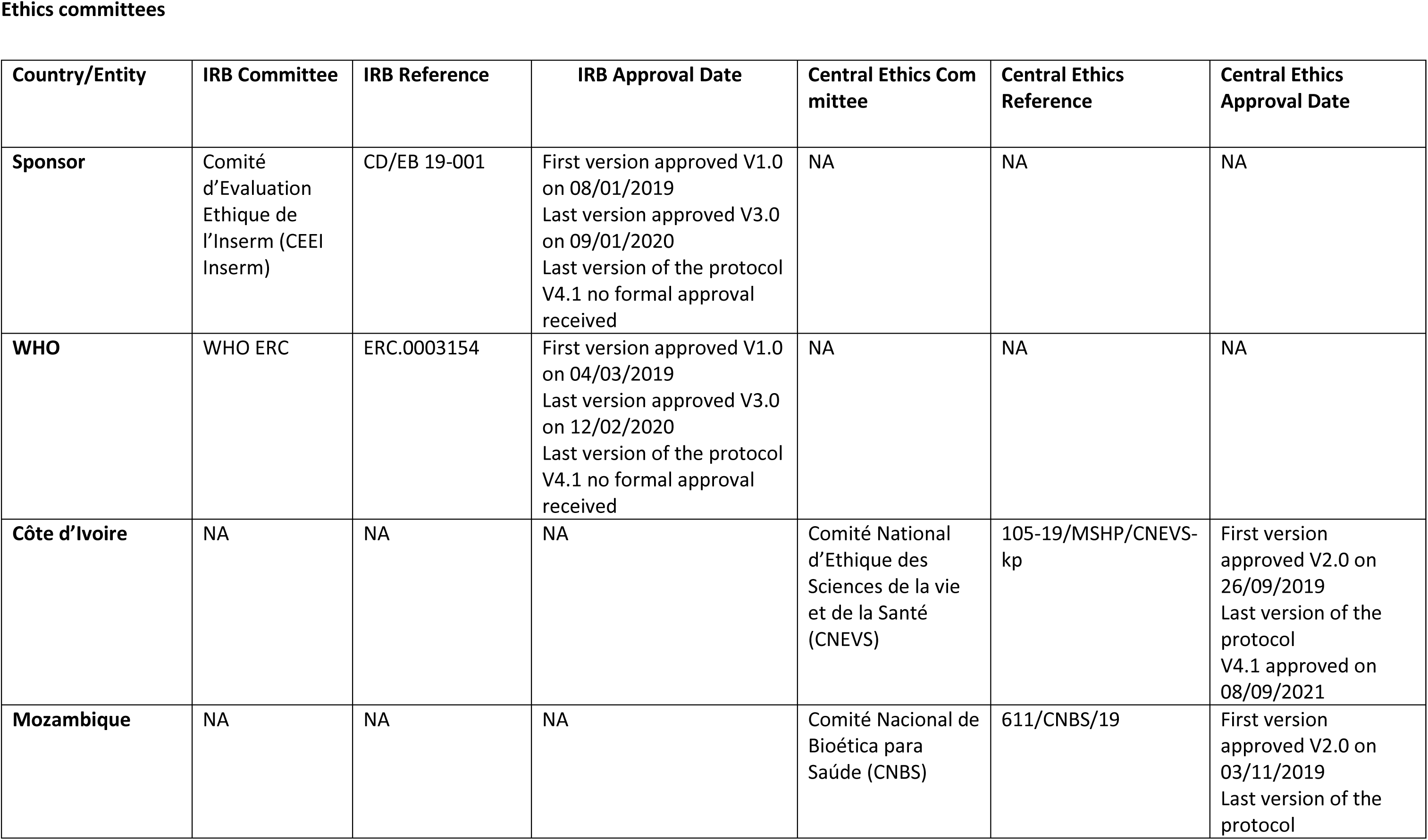
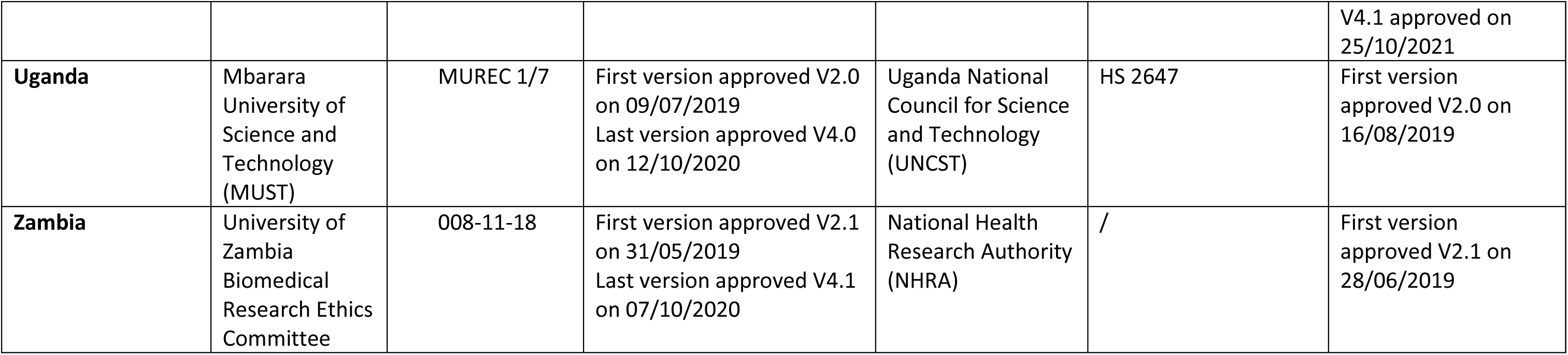

### Supplementary methods

Chest radiographies were performed with anteroposterior and lateral views in children aged ≤ 5 years and posteroanterior in older children. Additional abdominal ultrasonography features assessed included hepatomegaly, splenic or hepatic micro-abscesses, pericardial/pleural effusion, and/or ascites. Additional blood tests done included complete blood count (CBC), transaminases, C-reactive protein (CRP) and lymphocyte T-CD4 count were performed. The monocyte-lymphocyte ratio (MLR) was calculated. CBC, transaminases and CRP were monitored at M2 and M6 visit, and CD4 count at M6 visit.

The algorithm was considered completed if a decision to initiate TB treatment has been taken at any step of the algorithm or if TB has been excluded after systematic evaluation, and all steps planned in the algorithm have been implemented. Recent failure to thrive was defined as documented clear deviation from a previous growth trajectory in the last 3 months or Z score weight/age < 2.

As recommended per WHO 2011 guidelines, children not initiated on treatment as per PAANTHER TB-treatment decision algorithm could be initiated as soon as possible on TB preventive therapy ^27^. Antiretroviral treatment (ART) was started following WHO and national recommendations, i.e. 2 to 8 weeks of TB treatment initiation in children diagnosed with TB, and not later than 2 weeks in children with CD4 count below 50 cells/µL ^28^. In children already on ART, a change of antiretroviral regimen was considered if appropriate. Cotrimoxazole prophylactic treatments was prescribed according to the recommendations of national programs.

We hypothesized a prevalence of TB of 50%, resulting from a likely higher proportion of children on ART as compared to the PAANTHER study where the prevalence was 57.3% in CLHIV with presumptive TB as defined per standard algorithm entry criteria, with a proportion of 39.3% of children being on ART.

**Figure S1.**
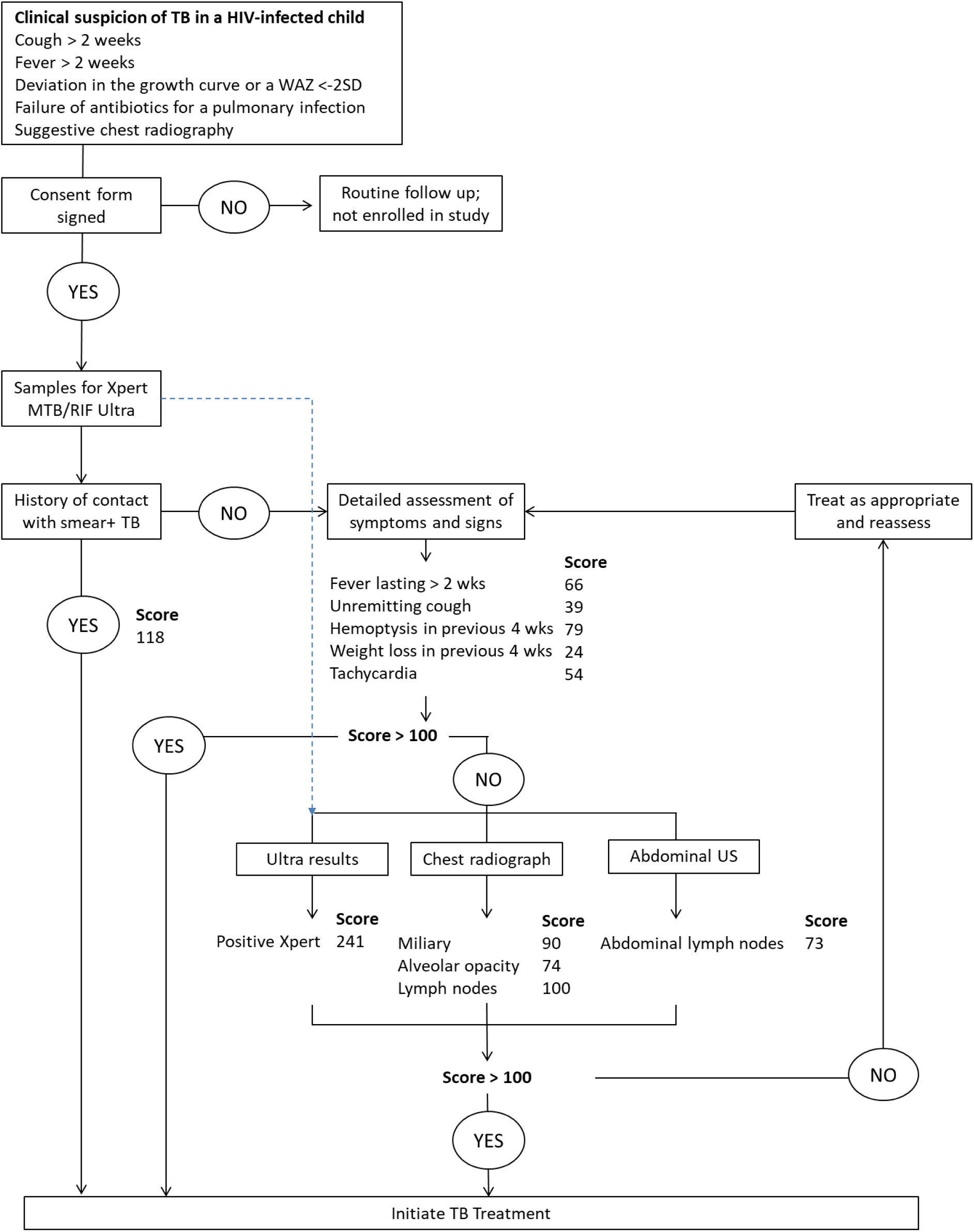
PAANTHER TDA.

**Figure S2.**
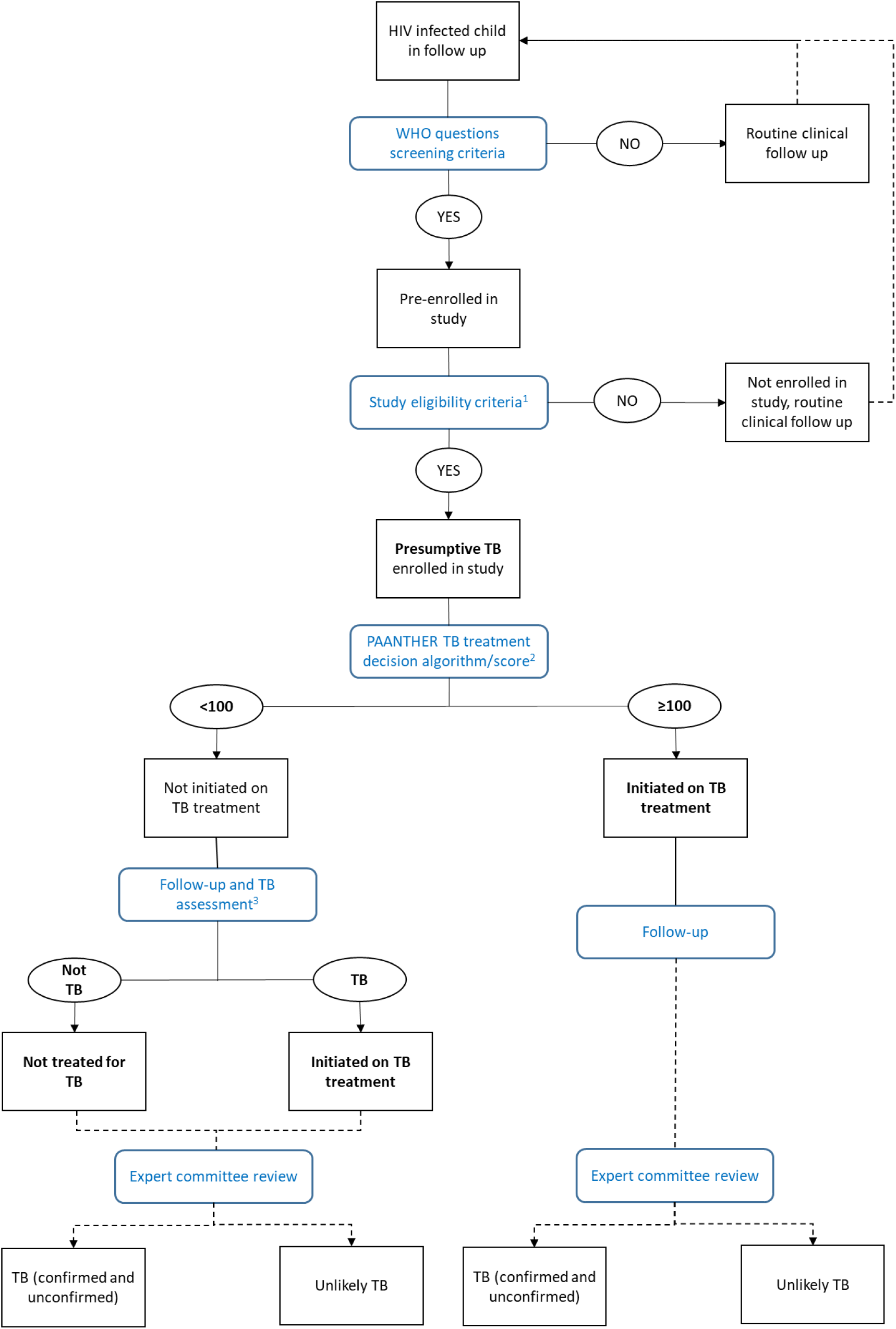
TB-Speed HIV study flow chart.

**Table S1:** PAANTHER TB treatment decision algorithm components and point scoring.

| Exam/assessment | Features/items | Points |
| --- | --- | --- |
| Contact history | History of contact with a smear + TB case | 118 |
| Symptoms | Fever lasting > 2 weeks | 66 |
|  | Unremitting cough | 39 |
|  | Hemoptysis in previous 4 weeks | 79 |
|  | Weight loss in previous 4 weeks | 24 |
| Clinical signs | Tachyardia | 54 |
| Xpert MTB/RIF Ultra | Positive Xpert MTB/RIF Ultra result | 241 |
| Chest radiography | Miliary | 90 |
|  | Alveolar opacity | 74 |
|  | Lymph nodes | 100 |
| Abdominal ultrasound | Abdominal lymph nodes | 73 |

**Table S2:** All children characteristics.

| Characteristic | N' if different from N | Overall N = 275<br>n (%) or median (IQR) | N' if different from N | Positive score N = 215<br>n (%) or median (IQR) | N' if different from N | Negative score N = 60<br>n (%) or median (IQR) | P-value |
| --- | --- | --- | --- | --- | --- | --- | --- |
| Age (years) |  | 5 (2·0; 10) |  | 5 (2·0; 10) |  | 5 (2·1; 10) | 0·98 |
| Sex Female |  | 142 (51·6) |  | 110 (51·2) |  | 32 (53·3) | 0·77 |
| Country |  |  |  |  |  |  | 0·031 |
| Ivory Coast |  | 26 (9·5) |  | 20 (9·3) |  | 6 (10·0) |  |
| Mozambique |  | 40 (14·5) |  | 28 (13·0) |  | 12 (20·0) |  |
| Uganda |  | 83 (30·2) |  | 74 (34·4) |  | 9 (15·0) |  |
| Zambia |  | 126 (45·8) |  | 93 (43·3) |  | 33 (55·0) |  |
| CD4 count (cells/mm3) | 208 | 736 (325·8; 1 302) | 165 | 730 (330·0; 1 283) | 43 | 870 (312·5; 1 424) | 0·51 |
| CD4 percentage | 206 | 21 [10·1; 30] | 164 | 21 [10·0; 29] | 42 | 20 [13·4; 30] | 0·94 |
| Immunosuppression* | 201 |  | 160 |  | 41 |  | 0·43 |
| Not significant |  | 75 (37·3) |  | 57 (35·6) |  | 18 (43·9) |  |
| Mild |  | 24 (11·9) |  | 22 (13·8) |  | 2 (4·9) |  |
| Advanced |  | 18 (9·0) |  | 15 (9·4) |  | 3 (7·3) |  |
| Severe |  | 84 (41·8) |  | 66 (41·2) |  | 18 (43·9) |  |
| On ART at inclusion | 250 | 175 (70·0) | 198 | 132 (66·7) | 52 | 43 (82·7) | 0·025 |
| Clinical history at baseline |  |  |  |  |  |  |  |
| Asthma |  | 2 (0·7) |  | 2 (0·9) |  | 0 (0·0) | >0·99 |
| Cardiac disorders |  | 3 (1·1) |  | 2 (0·9) |  | 1 (1·7) | 0·52 |
| Sickle cell disease |  | 6 (2·2) |  | 5 (2·3) |  | 1 (1·7) | >0·99 |
| BCG | 249 | 194 (70·5) | 192 | 150 (69·8) | 57 | 44 (73·3) | 0·41 |
| Previous TB treatment |  | 19 (6·9) |  | 14 (6·5) |  | 5 (8·3) | 0·74 |
| Pulmonary infection in the past month |  | 132 (48·0) |  | 117 (54·4) |  | 15 (25·0) | 0·47 |
| Failure of broad-spectrum antibiotics for a pneumonia | 274 | 61 (22·3) | 214 | 51 (23·8) |  | 10 (16·7) | 0·24 |
| History of contact with a smear or Xpert + tuberculosis case** | 274 | 18 (6·6%) |  | 17 (7·9%) | 59 | 1 (1·7) | 0·13 |
| Cough | 274 | 237 (86.5) |  | 196 (91.2) | 59 | 41 (69.5) | <0.0001 |
| Cough > 2 weeks | 274 | 172 (62.8) |  | 151 (70.2) | 59 | 21 (35.6) | <0.0001 |
| Unremitting cough** | 274 | 136 (49.6) |  | 119 (55.3) | 59 | 17 (28.8) | 0.0003 |
| Fever | 274 | 161 (58.8) |  | 138 (64.2) | 59 | 23 (39.0) | 0.0005 |
| Fever > 2 weeks** | 274 | 90 (32.8) |  | 84 (39.1) | 59 | 6 (10.2) | <0.0001 |
| Hemoptysis in previous 4 weeks** | 274 | 2 (0.7) |  | 2 (0.9) | 59 | 0 (0.0) | >0.99 |
| Weight loss in previous 4 weeks reported by parents/guardian** | 274 | 177 (64.6) |  | 142 (66.0) | 59 | 35 (59.3) | 0.34 |
| WAZ score | 201 | -3 (-4.1; -1) | 153 | -3 (-4.0; -1) | 45 | -3 (-4.3; -1) | 0.37 |
| SAM*** |  | 118 (42.9%) |  | 93 (43.3%) |  | 25 (41.7%) | 0.83 |
| Loss of appetite | 274 | 143 (52.2) |  | 117 (54.4) | 59 | 26 (44.1) | 0.16 |
| Fatigue or loss of playfulness | 274 | 140 (51.1) |  | 115 (53.5) | 59 | 25 (42.4) | 0.13 |
| Tachycardia** |  | 62 (22.5) |  | 53 (24.7) |  | 9 (15.0) | 0.11 |
| Cervical lymphadenopathy | 274 | 63 (23.0) |  | 55 (25.6) | 59 | 8 (13.6) | 0.052 |
| Hepatomegaly | 274 | 47 (17.2) |  | 42 (19.5) | 59 | 5 (8.5) | 0.046 |
| Chest radiography consistent with tuberculosis | 261 | 189 (72.4) | 205 | 176 (85.9) | 56 | 13 (23.2) | <0.0001 |
| Miliary** |  | 32 (12.3) |  | 31 (15.1) |  | 1 (1.8) | 0.0070 |
| Alveolar opacity** |  | 136 (52.1) |  | 126 (61.5) |  | 10 (17.9) | <0.0001 |
| Lymph nodes** |  | 126 (48.3) |  | 124 (60.5) |  | 2 (3.6) | <0.0001 |
| Cavitation |  | 25 (9.6) |  | 23 (11.2) |  | 2 (3.6) | 0.085 |
| Pleural effusion |  | 13 (5.0) |  | 12 (5.9) |  | 1 (1.8) | 0.31 |
| Airway compression |  | 4 (1.5) |  | 4 (2.0) |  | 0 (0.0) | 0.58 |
| Abdominal Lymph nodes** | 258 | 36 (14.0) | 204 | 35 (17.2) | 54 | 1 (1.9) | 0.0039 |
| Xpert MTB/RIF Ultra positive** | 271 | 25 (9.2) | 212 | 24 (11.3) | 59 | 1 (1.7)1 | 0.024 |
| Nasopharyngeal aspirate | 255 | 16 (6.3) | 201 | 15 (7.5) | 54 | 1 (1.9) | 0.21 |
| Stool | 241 | 13 (5.4) | 186 | 13 (7.0) | 55 | 0 (0.0) | 0.044 |
| Expectorated sputum | 56 | 2 (3.6) | 45 | 2 (4.4) | 11 | 0 (0.0) | >0.99 |
| Gastric aspirate | 201 | 7 (3.5) | 157 | 7 (4.5) | 44 | 0 (0.0) | 0.35 |
| MTB culture positive on any sample | 228 | 8 (3.5) | 179 | 8 (4.5) | 49 | 0 (0.0) | 0.21 |
| PAANTHER total score |  | 208 (118.5; 285) |  | 242 (179.0; 303) |  | 63 (39.0; 90) | <0.0001 |
| TB treatment initiated |  | 197 (71.6) |  | 185 (86.0) |  | 12 (20.0) | <0.0001 |
| Reference standard classification | 273 |  | 213 |  |  |  | <0.0001 |
| Confirmed |  | 25 (9.2) |  | 24 (11.3) |  | 1 (1.7) |  |
| Unconfirmed |  | 135 (49.5) |  | 127 (59.6) |  | 8 (13.3) |  |
| Unlikely |  | 113 (41.4) |  | 62 (29.1) |  | 51 (85.0) |  |
| End of study status |  |  |  |  |  |  | 0.24 |
| Alive |  | 198 (72.0) |  | 158 (73.5) |  | 40 (66.7) |  |
| Dead |  | 47 (17.1) |  | 37 (17.2) |  | 10 (16.7) |  |
| Transfer out |  | 1 (0.4) |  | 0 (0.0) |  | 1 (1.7) |  |
| Withdrawn |  | 12 (4.4) |  | 9 (4.2) |  | 3 (5.0) |  |
| Lost to follow-up |  | 17 (6.2) |  | 11 (5.1) |  | 6 (10.0) |  |

**Table S3.**
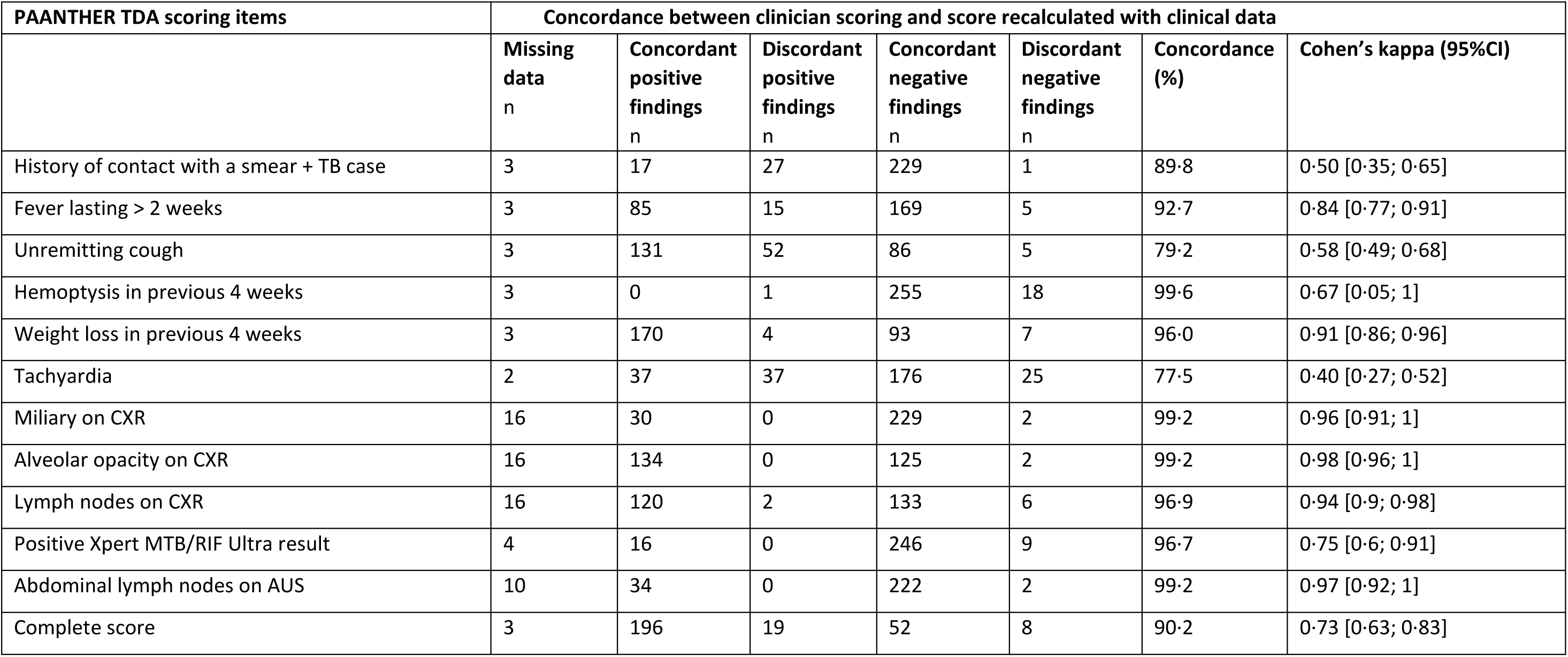
Concordance between clinician scoring and recalculated score.

| PAANTHER TDA scoring items | Concordance between clinician scoring and score recalculated with clinical data |  |  |  |  |  |  |
| --- | --- | --- | --- | --- | --- | --- | --- |
|  | Missing data<br>n | Concordant positive findings<br>n | Discordant positive findings<br>n | Concordant negative findings<br>n | Discordant negative findings<br>n | Concordance (%) | Cohen's kappa (95%CI) |
| History of contact with a smear + TB case | 3 | 17 | 27 | 229 | 1 | 89.8 | 0.50 [0.35; 0.65] |
| Fever lasting > 2 weeks | 3 | 85 | 15 | 169 | 5 | 92.7 | 0.84 [0.77; 0.91] |
| Unremitting cough | 3 | 131 | 52 | 86 | 5 | 79.2 | 0.58 [0.49; 0.68] |
| Hemoptysis in previous 4 weeks | 3 | 0 | 1 | 255 | 18 | 99.6 | 0.67 [0.05; 1] |
| Weight loss in previous 4 weeks | 3 | 170 | 4 | 93 | 7 | 96.0 | 0.91 [0.86; 0.96] |
| Tachyardia | 2 | 37 | 37 | 176 | 25 | 77.5 | 0.40 [0.27; 0.52] |
| Miliary on CXR | 16 | 30 | 0 | 229 | 2 | 99.2 | 0.96 [0.91; 1] |
| Alveolar opacity on CXR | 16 | 134 | 0 | 125 | 2 | 99.2 | 0.98 [0.96; 1] |
| Lymph nodes on CXR | 16 | 120 | 2 | 133 | 6 | 96.9 | 0.94 [0.9; 0.98] |
| Positive Xpert MTB/RIF Ultra result | 4 | 16 | 0 | 246 | 9 | 96.7 | 0.75 [0.6; 0.91] |
| Abdominal lymph nodes on AUS | 10 | 34 | 0 | 222 | 2 | 99.2 | 0.97 [0.92; 1] |
| Complete score | 3 | 196 | 19 | 52 | 8 | 90.2 | 0.73 [0.63; 0.83] |

**Table S4.**
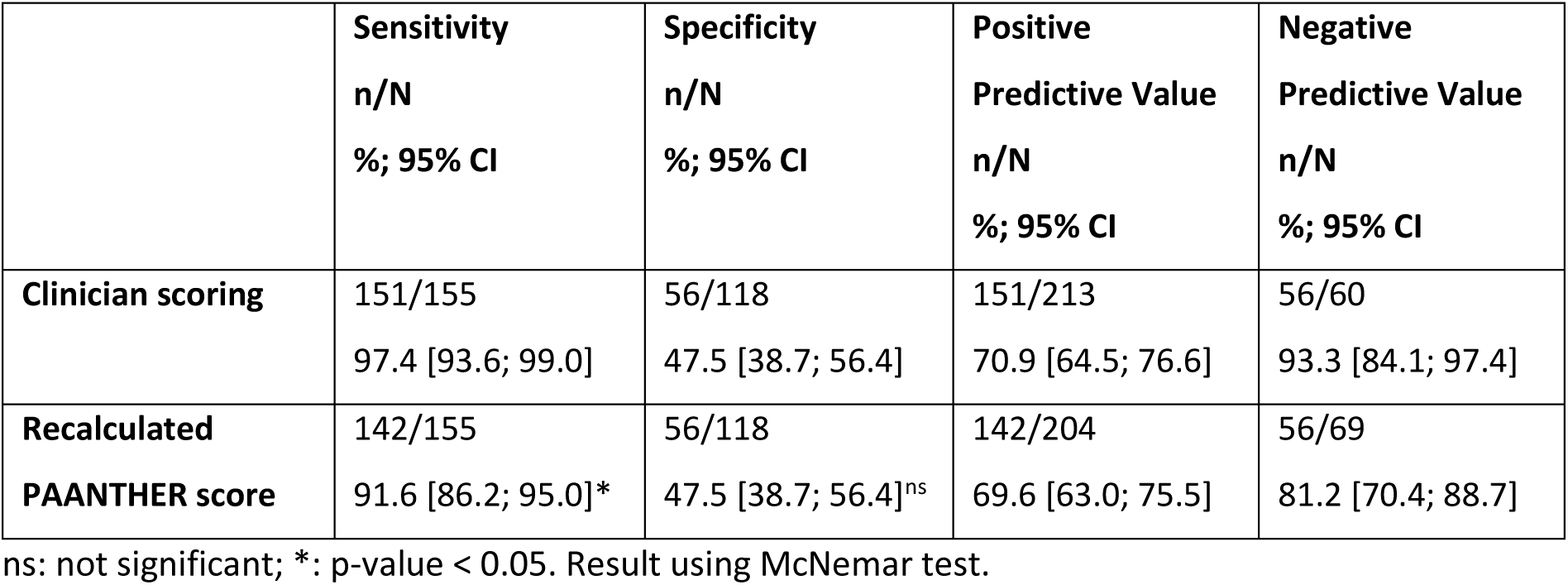
Sensitivity analyses of the PAANTHER score diagnostic performance.

**TB-Speed HIV Study Group**

**CÔTE D’IVOIRE**

**PACCI, Abidjan, Côte d’Ivoire:**

KOMENA Auguste Eric, MOH Raoul

**Mother and Child Department, Cocody University Teaching Hospital, Abidjan, Côte d’Ivoire:**

FOLQUET AMORRISSANI Madeleine

**Pediatrics Department, Angré University Teaching Hospital, Abidjan, Côte d’Ivoire**

Amon -Tanoh Dick

**Pediatrics Department, Treichville University Teaching Hospital, Abidjan, Côte d’Ivoire:**

CISSE Lassina

**FRANCE**

**University of Bordeaux, National Institute for Health and Medical Research (INSERM) UMR 1219, Research Institute for Sustainable Development (IRD) EMR 271, Bordeaux Population Health Centre, Bordeaux, France:**

BALESTRE Eric, BADRICHANI Anne, BEUSCART Aurélie, CHARPIN Aurélie, D’ELBEE Marc, FONT Hélène, HABIYAMBERE Gemma, HUYEN TON NU NGUYET Minh, MARCY Olivier, MESNIER Salomé, OCCELLI Estelle, POUBLAN Julien, RAZAFIMANANTSOA Manoa, ROUCHER Clémentine, SERRE Angeline, VESSIERE Aurélia

**University of Montpellier, IRD, INSERM, TRANSVIH MI, Montpellier, France:**

BONNET Maryline, CHAUVET Savine, LOUNNAS Manon

**National Institute for Health and Medical Research (INSERM), Paris, France:**

COUFFIN-CADIERGUES Sandrine, ESPEROU Hélène, HAMZE Benjamin, KUPPERS Alexis

**MOZAMBIQUE**

**Instituto Nacional de Saúde, Maputo, Mozambique:**

CASSY Sheyla, CHIÚLE Valter, CUMBE Cristina, CUMBE Saniata, GUNI Irene, KHOSA Celso, MACHONISSE Emelva, MATSHINHE Mércia, MILICE Denise, MUCHANGA Eva, RIBEIRO Jorge, ZITHA Alcina

**Ministry of Health, Maputo, Mozambique:**

JOSÉ Benedita, MANHÇA Ivan

**José Macamo General Hospital, Maputo, Mozambique:**

CAPATO Liliana, COMANDANTE Herquéria, FLORINDO Natércia, KITUNGWA Mule, MABOTA Roda, MACHEL Dália, MACHIANA Lídia, MENDES Helena, MUIANGA Valdo, REGO Dalila, TEMBE Luísa

**Maputo Central Hospital, Paediatrics Department, Maputo, Mozambique:**

ATUMANE Jafito, CHAVE Adélio, CHILUNDO Josina, FLORÊNCIO Marcelina, GIVE Josefina, LUCAS Gelson, MACASSA Eugénia, MAVALE Sandra, THAI Arsénio

**David Geffen School of Medicine, University of California, Los Angeles:**

**UGANDA**

**MUJHU Research Collaboration, MU-JHU Care Limited, Makerere University-John Hopkins University Research Collaboration,Kampala, Uganda**

WOBUDEYA Eric

**Epicentre Mbarara Research Centre, Mbarara, Uganda:**

KAKWENZA Paul, ARINAITWE Rinah, MWANGA-AMUMPAIRE Juliet, NATUKUNDA Naome

**Mbarara University of Science and Technology**

MWANGA-AMUMPAIRE Juliet, NASEJJE Milly

**Mbarara Regional Referral Hospital**

NASERA Denis, NAKIGOZI Patience

**Mulago National Referral Hospital, Kampala, Uganda:**

BABIREKERE Esther,

**Jinja Regional Referral Hospital, Jinja, Uganda**

MBEKEKA Prossy

**ZAMBIA**

**University Teaching Hospitals-Children’s Hospital (UTHC), Lusaka, Zambia - University of Zambia (UNZA), Lusaka, Zambia:**

CHABALA Chishala, MULENGA Veronica, CHIRWA Uzima, MUNDUNDU Gae KAPULA Chifunda, SHANKALALA Perfect, HAMBULO Chimuka, KAPOTWE Vincent, KASAKWA Kunda, CHILANGA Mutinta, CHILONGA Jessy, CHIMBINI Maria, CHOLA Daniel, MWANGO Eusters, NAWAKWI Grace, SIASULINGANA Teddy, ZULU Susan, HIMWAZE Diane

**Arthur Davidson Children’s Hospital, Ndola (ACDH), Zambia:**

CHAKOPO Moses, INAMBAO Muleya, NDUNA Bwendo

MUMBA Wyclef, MANKUNSHE Endreen, PUMBWE Mwamba, HALENDE Barbara, MOONO Roy, SILAVWE Maureen

**Endpoint review committee**

CHUNGU Chalilwe, ZIMBA Kevin, ZYAMBO Khozya, CHIRWA Uzima, KAPASA Monica

**Study monitors**

KANYAMA Mirriam, NGAMBI Marjory

## Notes

### Competing Interest Statement

The authors have declared no competing interest.

### Clinical Trial

NCT04121026

### Clinical Protocols

https://www.tb-speed.com/wp-content/uploads/2022/08/TB-Speed-HIV-Study-Protocol.pdf

### Funding Statement

This study was funded by UNITAID

### Author Declarations

The IRB of the Comité d'Évaluation & Éthique de l'Inserm (CEEI Inserm) granted ethical approval for this work. The IRB of the WHO ERC granted ethical approval for this work. The Central Ethics Committee of the Comité National d'Éthique des Sciences de la Vie et de la Santé (CNEVS), Côte d'Ivoire, granted ethical approval for this work. The Central Ethics Committee - Comité Nacional de Bioética para a Saúde (CNBS), Mozambique, granted ethical approval for this work. The IRB of Mbarara University of Science and Technology (MUST) granted ethical approval for this work. The Central Ethics Committee of the Uganda National Council for Science and Technology (UNCST) granted ethical approval for this work. The IRB of the University of Zambia Biomedical Research Ethics Committee granted ethical approval for this work. The Central Ethics Committee of the National Health Research Authority (NHRA), Zambia, granted ethical approval for this work.

